# Clinical Characteristics of 34 Children with Coronavirus Disease-2019 in the West of China: a Multiple-center Case Series

**DOI:** 10.1101/2020.03.12.20034686

**Authors:** Che Zhang, Jiaowei Gu, Quanjing Chen, Na Deng, Jingfeng Li, Li Huang, Xihui Zhou

## Abstract

**Background:** Up to 9 March, 2020, 109577 patients were diagnosed with coronavirus disease-2019 (COVID-19) globally. The clinical and epidemiological characteristics of adult patients have been revealed recently. However, the information of paediatric patients remains unclear. We describe the clinical and epidemiological characteristics of paediatric patients to provide valuable insight into early diagnosis of COVID-19 in children, as well as epidemic control policy making.

**Methods and Findings:** This retrospective, observational study was a case series performed at 4 hospitals in the west of China. Thirty-four paediatric patients with COVID-19 were included from January 1 to February 25, 2020. And the final follow-up visit was completed by February 28, 2020. Clinical and epidemiological characteristics were analyzed on the basis of demographic data, medical history, laboratory tests, radiological findings, and treatment information. Data analysis was performed on 34 paediatrics patients with COVID-19 aged from 1 to 144 months (median 33.00, IQR 10.00 - 94.25), among whom 14 males (41.18%) were included. 47.60% of patients were noticed without any exposure history. The median incubation period was 10.50 (7.75 - 25.25) days. Infections of other respiratory pathogens were reported in 16 patients (47.06%). The most common initial symptoms were fever (76.47%), cough (58.82%), and expectoration (20.59%). Vomiting (11.76%) and diarrhea (11.76%) were also reported in a considerable portion of cases. A remarkable increase was detected in serum amyloid A for 17 patients (85.00%) and high-sensitivity C-reactive protein for 17 patients (58.62%), while a decrease of prealbumin was noticed in 25 patients (78.13%). In addition, the levels of lactate dehydrogenase was increased significantly in 28 patients (82.35%), as well as α-hydroxybutyrate dehydrogenase in 25 patients (73.53%). Patchy lesions in lobules were detected by chest computed tomographic scans in 28 patients (82.36%). The typical feature of ground-glass opacity for adults was rare in paediatric patients (2.94%). A late-onset pattern of lesions in lobules were also noticed. Stratified analysis of the clinical features were not performed due to relatively limited samples.

**Conclusions:** Our data presented the clinical and epidemiological features of paediatric patients systemically. The findings offer new insight into the early identification and intervention of paediatric patients with COVID-19.

**Author summary:** *Why was this study done?:*

- The 2019-novel coronavirus (2019-nCoV) infection has spread worldwide rapidly.
- Early identification and intervention are necessary for effective epidemic control in both adults and children, however the clinical and epidemiological characteristics of paediatric patients remains unclear.

*What did the researchers do and find?:*

- We collected and analyzed clinical data of 34 paediatric patients with coronavirus disease-2019 (COVID-19) in 4 hospitals of China from January 1 to February 25, 2020.
- We described the clinical and epidemiological features of the patients, and focused on the differences in initial symptoms and radiological findings between paediatric patients and adult patients.
- Distinguished from adult patients, higher incidences of fever, vomiting, and diarrhea were noticed on admission in paediatric cases.
- Patchy shadows of high density were common in lobules lesions, while the typical features of ground-glass opacity in adults were rare in paediatric cases.
- A late-onset pattern of lobules lesions was revealed on the basis of chest computed tomographic scans.

*What do these findings mean?:*

- The specific clinical features in paediatric patients should be paid more attention to, when the physicians are dealing with suspected cases.
- The epidemiological model in children was characterized with dominant family cluster transmission and extended incubation period, which should be taken into consideration in policy making for epidemic control.

## Introduction

The 2019-novel coronavirus (2019-nCoV) infection has spread worldwide rapidly since its outbreak from Wuhan city in China in early December 2019 [1]. The epidemic of 2019-nCoV has been declared as a public health emergency of international concern by World Health Organization (WHO) on January 30, 2020 [2]. Up to date, 80904 cumulative laboratory-confirmed cases have been reported in China. Meanwhile, 28673 cases have been reported from 104 countries and regions outside of China [3]. The 2019-nCoV was identified as a diverse clade from the severe acute respiratory syndrome coronavirus (SARS-CoV) and Middle East respiratory syndrome coronavirus (MERS-CoV), and was reported as the cause of coronavirus disease-2019 (COVID-19) [4]. The clinical characteristics of adult patients with COVID-19 have been revealed in recent studies, mainly including fever, cough, dyspnea, and radiographic findings of pneumonia [5-7]. However, the information of paediatric patients remains unclear.

This case series describes the clinical and epidemiological features of 34 paediatric patients on the basis of epidemiological, demographic, laboratory as well as radiological data, and aims to contribute to a comprehensive understanding of the characteristics of COVID-19.

## Methods

### Study Design and Participants

This retrospective, observational study was approved by the institutional review board (IRB) of Affiliated Taihe Hospital of Hubei University of Medicine (Ethical approval No.2020KY01). Suspected patients with clinical and/ or radiological features of pneumonia were isolated for 2019-nCoV nucleic acid detection according to relevant guidelines [8, 9]. Admitted children with laboratory-confirmed 2019-nCoV positive results were included in the study from 4 hospitals of China during January 1 to February 23, 2020. And the final follow-up visit was completed by February 28, 2020. Informed consent was obtained from the patients and their guardians prior to data collection.

### Laboratory detection for 2019-nCoV

Samples for pathogen detection were obtained from throat swab or lower respiratory tract specimen. Real-time reverse transcription polymerase chain reaction (RT-PCR) was employed for 2019-nCoV detection. Briefly, RNA was extracted from samples using RNA isolation kit (SDK60103, bioPerfectus technologies, Taizhou, China) according to the manufacturer’s instructions. After lysis and centrifugation, the suspension was collected for 2019-nCoV nucleic acid detection. RT-PCR assay was preformed using 2019-nCoV specific kit (NC-ORF1ab/N, DAAN GENE, Guangzhou, China). The adapter primers and probe were supplied with kits, targeting to the open reading frame 1ab (*ORF1ab*) and nucleocapsid protein (*N*) of CoV gene in accordance with the recommendation of National Health Commission of the People’s Republic of China [10]. Specifically, for target *ORF1ab*: forward primer sequence CCCTGTGGGTTTTACACTTAA; reverse primer sequence ACGATTGTGCATCAGCTGA; and the probe sequence 5’-FAM-CCGTCTGCGGTATGTGGAAAGGTTATGG-BHQ1-3’. For target *N*: forward primer sequence GGGGAACTTCTCCTGCTAGAAT; reverse primer sequence CAGACATTTTGCTCTCAAGCTG; and the probe sequence 5’-FAM-TTGCTGCTGCTTGACAGATT-TAMRA-3’. Amplifications were performed according to manufacturer’s protocol under following conditions: reverse transcription at 50°C for 15 min, and Taq inhibitor inactivation at 95°C for 15 min, followed by 45 cycles of denaturation at 94°C for 15 s, and extension at 55°C for 45 s. The results of 2019-nCoV nucleic acid detection were analyzed following the manufacturer’s instructions. Specifically, a test result with a cycle threshold value (Ct-value) ≤ 38 and a significant amplification curve was defined as positive. And a negative result was defined with a Ct-value > 38. Retesting was required in cases other than those mentioned above. Besides, negative cases were double checked by re-sampling and retesting with an interval as 24 h, and could be confirmed when negative results were obtained in 2 consecutive tests.

### Data Collection

The medical records of included patients were accessed by study team for data collection. And the data was cross-checked by staff from the Institute of Drug Clinical Trial of Taihe Hospital for quality control. Specifically, demographic information, medical history, and exposure history were collected from patients and their guardians. Vital signs, physical exams, laboratory tests, and electroencephalogram (ECG) were performed and recorded in routine process for inpatient paediatric patients. Chest computed tomographic (CT) scans were performed to detect signs of pneumonia if any. In particular, other respiratory pathogens were detected for differential diagnosis screening, including influenza A virus, influenza B virus, parainfluenza virus, respiratory syncytial virus, adenovirus, epstein-barr virus, and mycoplasma pneumoniae. Therapeutic information was also collected from electronic medical records. In addition, the median incubation period and disease course were analysed for epidemiological and clinical features of paediatric patients with COVID-19.

### Statistical Analysis

Descriptive statistics were performed using SPSS software (version 20.0, IBM, Armonk, NY, USA). No imputation was made for missing data. Categorical variables are presented as number and frequency rates, and continuous variables are presented as median and interquartile range (IQR).

## Results

### Characteristics of the Patients

In this case series, data from 34 paediatric patients with COVID-19 were analyzed, including 14 male patients (41.18%) and 20 female patients (58.82%). The median age was 33 (IQR 10.00 - 94.25) months with a range of 1 to 144 months. Eighteen patients (52.40%) had exposure to residents of Wuhan, among whom history of close contact was reported in 13 patients (72.22%). In particular, infections of other respiratory pathogens were reported in 16 patients (47.06%), including mycoplasma pneumoniae (26.47%), influenza B virus (17.65%), influenza A virus (8.82%), respiratory syncytial virus (5.88%), epstein-barr virus (5.88%), parainfluenza virus (2.94%), and adenovirus (2.94%). Comorbidities were reported in 6 patients (17.65%). In respect of initial symptoms and signs, fever (76.47%), cough (58.82%), and expectoration (20.59%) were complained most frequently. Meanwhile, tachypnea, vomiting, and diarrhea were reported as well (Table 1). Regarding the disease course, the median incubation period was 10.50 (7.75 - 25.25) days. Moreover, the median day was 2.00 (1.00 - 3.50) from symptom onset to admission, and 3.00 (2.00 - 4.00) between admission and recover from fever (Fig. 1).

**Table 1.** Demographics and Characteristics of Patients on Admission.

|  | Data at baseline (*n* = 34) |
| --- | --- |
| Demographics |  |
| Sex |  |
| Male, *n* (%) | 14 (41.18) |
| Female, *n* (%) | 20 (58.82) |
| Age, median (IQR), m | 33.00 (10.00 - 94.25) |
| History of exposure, *n* (%) | 18 (52.40) |
| Viral infection other than 2019-nCoV, *n* (%) | 16 (47.06) |
| Comorbidities |  |
| Infectious Mononucleosis, *n* (%) | 2 (5.88) |
| Nephroblastoma, *n* (%) | 1 (2.94) |
| Atrial septal defect, *n* (%) | 1 (2.94) |
| Febrile convulsion, *n* (%) | 1 (2.94) |
| Asthma, *n* (%) | 1 (2.94) |
| Signs and symptoms |  |
| Fever, *n* (%) | 26 (76.47) |
| Cough, *n* (%) | 20 (58.82) |
| Expectoration, *n* (%) | 7 (20.59) |
| Vomiting, *n* (%) | 4 (11.76) |
| Diarrhea, *n* (%) | 4 (11.76) |
| Tachypnea, *n* (%) | 3 (8.82) |
| Intervention before admission, *n* (%) | 30 (88.24) |

Abbreviations: 2019-nCoV, 2019 novel coronavirus; IQR, interquartile range

**Fig 1.**
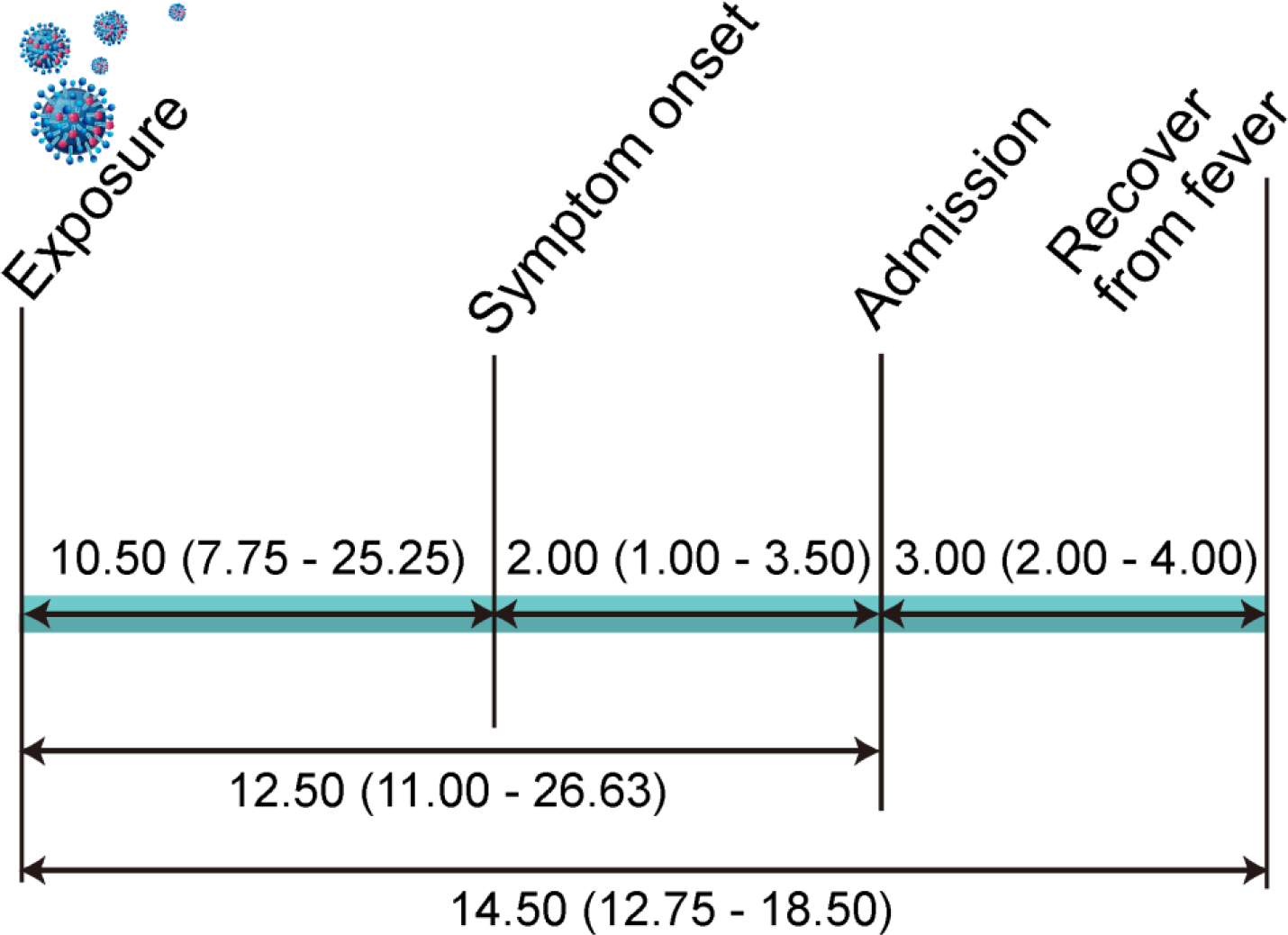
Duration of COVID-19 in Paediatric Patients. The timeline of disease course was presented as median (IQR) days. Incubation period was defined as the interval from the potential earliest date of exposure to the transmission source (residents of Wuhan) and the initial symptom onset date. Abbreviations: IQR, interquartile range

### Laboratory Tests Findings and Treatments

On admission, hematology tests indicated lymphocyte count was increased in 17 patients (50.00%), although the median value (3.19, 1.73 - 4.34) was shown within normal range. Concerning the findings in blood biochemistry, prealbumin (median 138.65 mg /L) decreased significantly in 25 patients (78.13%), while a remarkable increase was detected in serum amyloid A (SAA) for 17 patients (85.00%) and high-sensitivity C-reactive protein (hs-CRP) for 17 patients (58.62%). In addition, a noticeable increase was observed in lactate dehydrogenase (LDH) for 28 patients (82.35%) and α-hydroxybutyrate dehydrogenase (α-HBDH) for 25 patients (73.53%). However, the results of creatine kinase (CK) and creatine kinase-MB (CK-MB) were in normal level for all patients (Table 2), as well as ECG exam. No other significant findings were observed in blood coagulation tests, urine and stool routine tests.

**Table 2.**
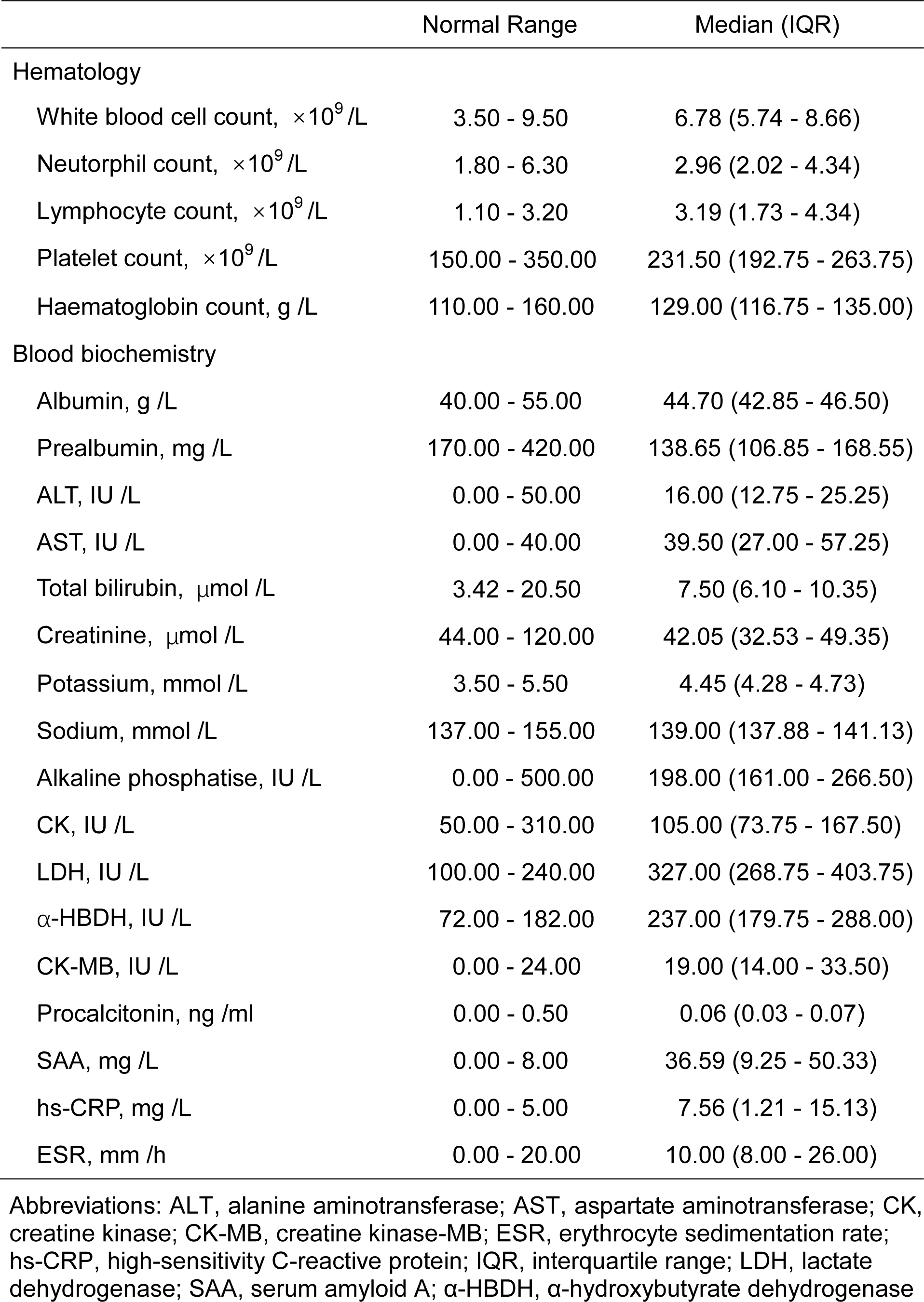
Laboratory Findings of Patients on Admission (*n*=34)

Multimodality therapies were employed, including antibiotic therapy (88.24%), antiviral therapy (82.35%), interferon-α nebulization (82.35%), corticosteroid therapy (14.71%), and oxygen inhalation supportive therapy (8.82%) (Table 3). Table 3. Chest Computed Tomographic Images Finding and Treatments of Patients

**Table 3.**
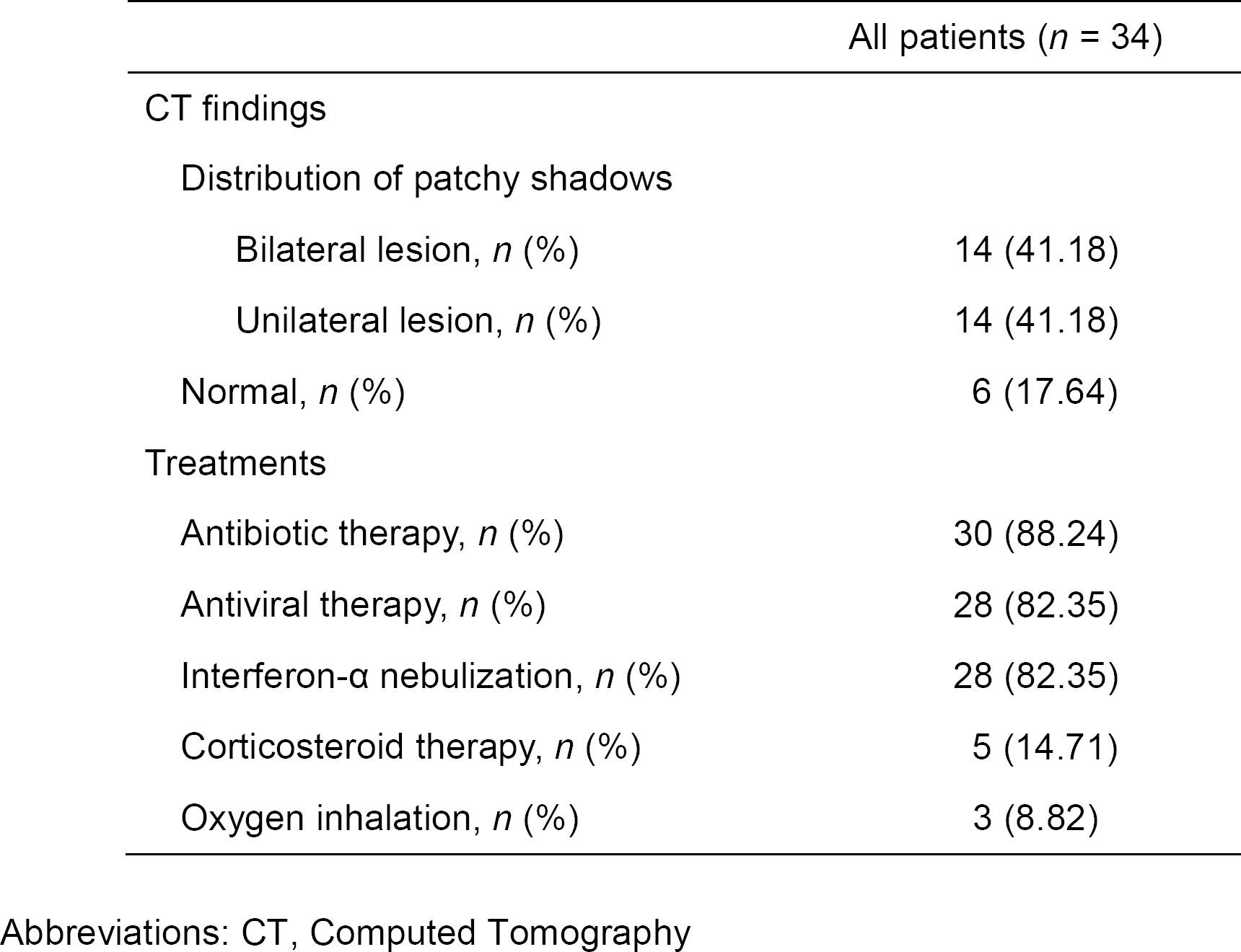
Chest Computed Tomographic Images Finding and Treatments of Patients.

### Chest CT Images Findings

Patchy lesions in lung lobules were indicated by chest CT scans in 28 patients (82.36%) on admission. However, lesions in lobules were detected in 4 - 5 days after admission for 5 cases (14.71%) among the patients with normal initial CT image on admission. There was only 1 case (2.94%) without abnormal radiological finding during hospitalization. Moreover, unilateral lesions were detected in 14 patients (41.18%), while bilateral lesions were shown in 14 patients (41.18%) (Table 3). The typical features of lesions were presented with patchy shadows of high density in lobules (Fig 2).

**Fig 2.**
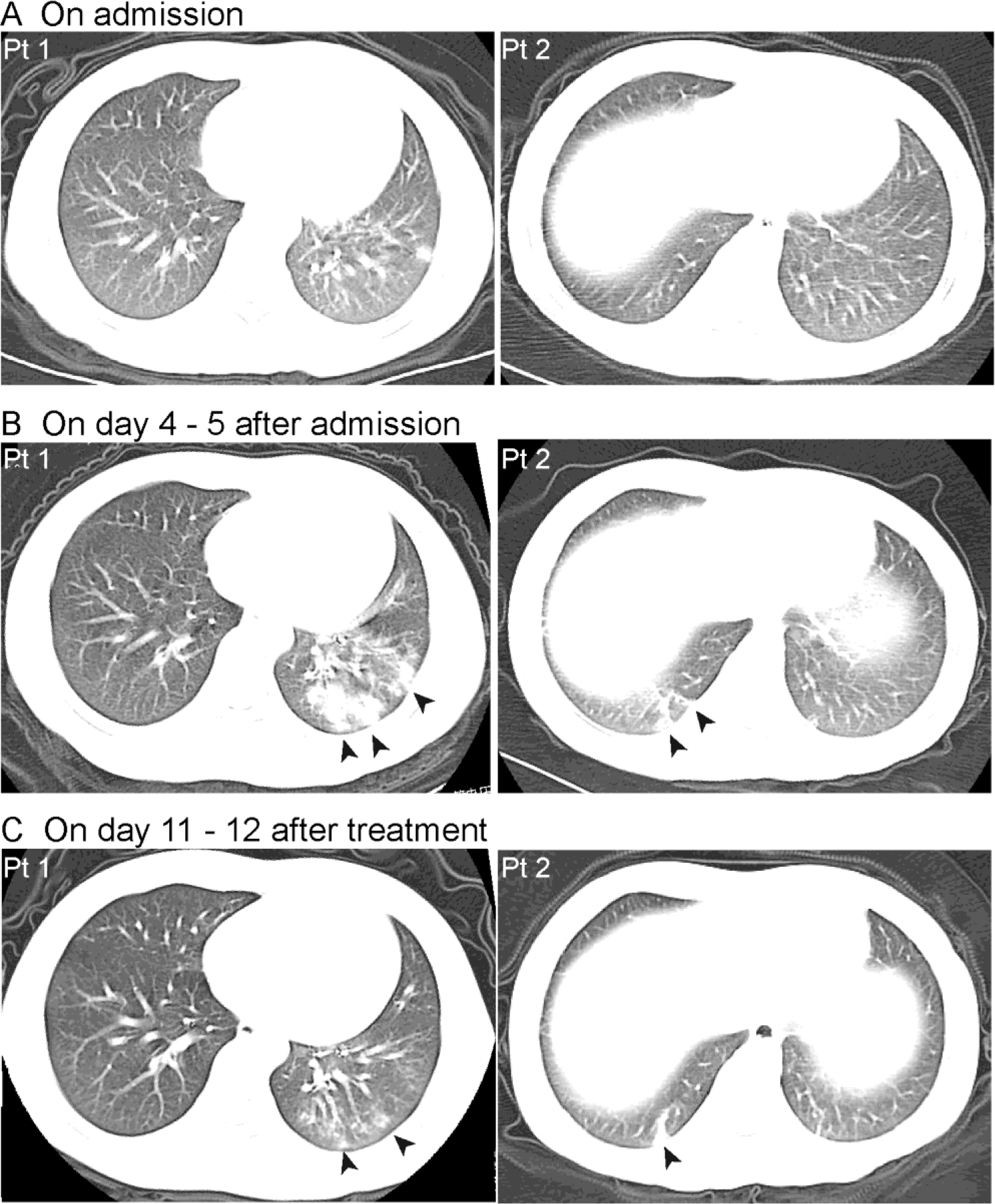
Typical Features of CT Images of Paediatric Patients with COVID-19. A. Initial CT images were obtained on admission without abnormal findings for Pt 1 (on February 1, 2020) and for Pt 2 (on February 5, 2020). B. The lesions in lobules were indicated by patchy shadows of high density for Pt 1 (on February 6, 2020, day 5 after admission) and Pt 2 (on February 9, 2020, day 4 after admission). C. The lesions in lobules still existed in discharge of Pt 1 (on February 12, 2020, day 12 after treatment) and Pt 2 (on February 15, 2020, day 11 after treatment). Abbreviations: CT, computed tomographic; Pt, patient 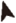 patchy shadows of high density in lobules

## Discussion

As mentioned in the literature review, the morbidity of COVID-19 was reported as 0.9% among children age 0 - 14 [1]. However, the clinical and epidemiological characteristics of paediatric patients haven’t been determined clearly yet. So far, this is the largest case series to present the clinical and epidemiological characteristics in children with COVID-19, as well as the first study to analyze the clinical features in paediatric patients systematically.

Our findings in paediatric patients indicated several features distinguished from adult patients, concerning epidemiological data, characteristics, initial symptoms, laboratory tests, and CT findings. In respect to exposure history, up to 72.30% of non-residents of Wuhan had contact with residents of Wuhan [1]. Thus, close contact with residents of Wuhan was indicated as a high risk factor for transmission in adult patients. However, a considerable proportion of patients (47.60%) were noticed without any exposure history in our study. The unanticipated finding suggested the referential value of exposure history for diagnosis of COVID-19 should be considered carefully for paediatric patients. In addition, family cluster transmission was common in paediatric patients. Close dynamic monitoring for families with adult patients would contribute to early identification for paediatric patients. In accordance with the features of indirect contact exposure, the incubation period was 10.50 (7.75 - 25.25) days in paediatric patients, while 4.00 (2.00 - 7.00) days of incubation was revealed for patients in all age groups [1]. The epidemiological features of paediatric patients indicated dynamic observation was necessary for suspected cases in children due to extended incubation period.

Dramatically, infections of other respiratory pathogens were detected in 16 patients (47.06%), however negative results were shown for these pathogens in adult patients [6]. In alignment with the results of pathogens detection, a response to viral infection was indicted by laboratory tests. Increasing lymphocyte count was detected in 17 (50.00%) patients. The levels of SAA and hs-CRP were increased. Meanwhile, decreasing level of prealbumin was noticed.

Basically, mild cases were common in paediatric patients with COVID-19 [1], presenting with acute upper respiratory tract infection or mild pneumonia. Generally, comorbidities were considered to predict the clinical outcomes. In our study, comorbidities were found in 6 patients (17.65%), similar to that in adult patients with mild symptoms (21.00%) [1]. In addition, effective control of disease was attributed to the prompt access to medical care in paediatric patients. Intervention before admission was reported in 30 patients (88.24%) upon initial symptom onset. And the median duration was 2.00 (1.00 - 3.50) days from symptom onset to admission. Typically, initial symptoms were valuable to predict the suspected cases. The most common initial symptom, fever, was identified in 26 children (76.47%) in our study, however it was presented in only 43.8% of adults patients on admission [1]. Besides, vomiting (11.76%) and diarrhea (11.76%) were also complained on admission, which were more common than that in adult patients (5.00% for vomiting and 3.80% for diarrhea) [1]. Consistent with previous reports for adults [1, 6], the levels of LDH and α-HBDH were increased remarkably without any symptom or sign of myocardial impairment.

According to our current data in chest CT images, patchy shadows were detected in 82.36% of patients on admission, in accordance with that in adults (86.20%) [1]. We also noticed the lesions in lobules were characterized with patchy shadows of high density in most cases (97.06%). Meanwhile, the typical feature of ground-glass opacity in adults (56.4%) [1] was rare in paediatric patients (2.94%). It was noteworthy that lesions in lobules would develop promptly in a few days, since they could be shown initially in some cases (14.71%) in 4 - 5 days after admission. The late-onset pattern of lesions was indicated in initial CT scan after 6 - 12 days upon symptom onset [11]. However, the clinical presentations were not so aggressive as the signs shown in CT images. Nonetheless, our findings suggested a close monitoring for paediatric patients due to the aggressive development of lesions in lobules, and the late-onset pattern in some cases as well.

The therapeutic strategy was on the basis of antiviral and antibiotic therapies, which was in alignment with the recommendations of National Health Commission of the People’s Republic of China [12]. The patients recovered from fever in 3.00 (2.00 - 4.00) days upon admission after treatments. The lesions in lobules still existed in discharge of patients, although great improvements were shown in CT scans after treatments.

The current study was limited by small sample size due to low morbidity in paediatric patients. The small sample size did not allow for intensive epidemiological analysis and stratified analysis of the clinical features. Further information in this field would help us to figure out a whole picture of COVID-19.

## Conclusion

This case series described the clinical and epidemiological characteristics in paediatric patients with COVID-19. Our data presented the clinical features of paediatric patients to facilitate early identification and intervention of suspected patients. Notwithstanding the relatively limited samples, our findings offer valuable insight into early diagnosis and epidemic control of COVID-19 in children.

## Data Availability

Individual participant data that underlie the results reported in this article are available after deidentification for investigational purpose. Data are available in our hospital's clinical trial medical records managed by GCP office with investigator’s support.

## Acknowledgments

None.

